# Knowledge and practices toward occupational health and environmental sustainability among farmers in Pabna District, Bangladesh: a cross-sectional study

**DOI:** 10.64898/2026.10.04.26364727

**Authors:** Shahnaj Parvin, Md. Obaidur Rashid, Md. Maksudur Rahman, Abdullah Al Zaber, Md. Bakhtiar Hossain, Rokeya Begum, Md. Azizul Haque

## Abstract

Bangladesh’s agricultural sector remains heavily dependent on pesticides and chemical inputs, yet limited attention has been paid to how farmers’ knowledge of occupational health and environmental sustainability translates into actual practice across the full agricultural value chain. This study aimed to assess the knowledge and practices of farmers in Pabna District, Bangladesh, regarding occupational health, pesticide handling, environmentally sustainable farming, and market sanitation. A cross-sectional survey was conducted among 301 farmers in 2024 using a structured questionnaire covering socio-demographic characteristics, sustainable farming knowledge and adoption, soil testing and organic farming practices, pesticide handling and occupational health and safety behaviours, post-harvest waste management, and market sanitation conditions. Data were analysed using descriptive statistics. The majority of respondents were male (87.04%) and had no formal education (63.46%). Although most farmers reported knowledge of environmentally sustainable farming practices (97.70%) and pesticide label instructions (98.00%), corresponding safety practices were poor: only 10.63% reported wearing any personal protective equipment and 1.33% reported using a mask during pesticide application. Eye irritation was the most commonly self-reported health symptom following pesticide exposure (92.69%). Market sanitation infrastructure was similarly inadequate, with 89.00% of respondents reporting an absence of sanitary toilets. These findings indicate a consistent gap between farmers’ knowledge and their actual practices across multiple domains of the agri-food system. Strengthening extension services, delivering practical hands-on occupational safety training, and investing in market sanitation infrastructure are recommended to close this gap and support safer, more sustainable farming practices in Bangladesh.

## 1. Introduction

Bangladesh is a densely populated country in South Asia, with a population of approximately 159 million and a population density of around 1,237 people per km² [1]. The country depends heavily on agriculture, which employs nearly 43% of the working population and contributes a significant share of gross domestic product (GDP) [2]. To meet growing food demand, Bangladeshi farmers rely extensively on pesticides and chemical fertilisers to protect crops and increase yield [3, 4]. Pesticides are widely recognised as poisonous and hazardous materials when handled unsafely [5] and studies conducted across developing countries have repeatedly shown that farmers’ pesticide practices are often unsafe, resulting in a wide range of health problems, including gastrointestinal, dermatological, and reproductive effects [6], as well as neurological and carcinogenic effects in cases of prolonged exposure [7, 8]. In Bangladesh specifically, organophosphate compounds account for the majority of reported pesticide poisoning cases [9], and unsafe practices such as smoking during pesticide application or failing to use protective equipment are common contributors to this risk [10].

Interestingly, evidence from several countries suggests that the relationship between farmers’ knowledge and their actual practice is not always linear; simply knowing about pesticide hazards is often not enough to change behaviour. This gap has been documented directly in several studies. In Kuwait, Jallow et al. (2017) found that although most farmers understood pesticide hazard information, the majority still did not use any personal protective equipment when mixing or spraying [11]. Similarly, Istriningsih et al. (2022) found that Indonesian farmers demonstrated reasonably good knowledge of Good Agricultural Practices (GAP) for safe pesticide use, yet showed markedly lower levels of actual application, particularly regarding the use of gloves, masks, and proper container disposal [12]. These findings indicate that knowledge alone is insufficient to ensure safe practice. This knowledge-practice gap is not confined to pesticide handling alone: similar patterns have been observed in related areas of agricultural sustainability. Soil testing and fertiliser recommendation services remain underused in Bangladesh despite their availability [13, 14], and farmers frequently apply cow dung to their fields without a structured understanding of organic farming principles [15, 16]. Beyond the farm itself, agricultural markets in many developing countries commonly lack adequate waste disposal and sanitation infrastructure despite general awareness of proper waste management practices [17].

Although the knowledge-practice gap in pesticide safety has received some research attention, most existing studies examine a single domain in isolation, typically pesticide handling alone, with few studies assessing this gap comprehensively across the full range of practices that shape farmer health and environmental sustainability within a single farming population. This is particularly true for Bangladesh, where no study to date has examined this gap across pre-harvest, pesticide handling, and post-harvest/market domains together. The present study aims to address this gap. It was conducted in Pabna District, located in the Rajshahi Division of north-western Bangladesh, a district known for cultivating rice, wheat, jute, mustard, onion, and brinjal, where agriculture remains the principal source of income for most residents [18]. Like many agricultural regions of Bangladesh, farming in Pabna relies heavily on chemical inputs, making it an appropriate setting in which to examine the relationship between farmers’ knowledge and practices related to occupational health and environmental sustainability.

Accordingly, this study sought to answer the following research question: to what extent does farmers’ knowledge of occupational health and environmental sustainability align with their actual practices across the agricultural value chain in Pabna District, Bangladesh? Specifically, this study aimed to examine, among farmers in Pabna District: (i) their socio-demographic characteristics; (ii) their knowledge and adoption of environmentally sustainable farming practices; (iii) their knowledge and practices regarding soil testing, organic farming, and manure use; (iv) their pesticide handling practices and occupational health and safety behaviours; (v) their post-harvest waste management practices and local market sanitation conditions; and (vi) health symptoms self-reported following pesticide exposure. The findings of this study are expected to inform more effective training and extension programmes for farmers, and to support policy efforts aimed at closing the gap between agricultural knowledge and safe, sustainable practice in Bangladesh.

## 2. Methods

### 2.1 Study design and setting

A cross-sectional study was conducted between July 2024 and August 2024 to assess farmers’ knowledge and practices related to occupational health, pesticide management, environmental sustainability, and market sanitation in Pabna District, Bangladesh.

Pabna District is located in northern Bangladesh (24.006°N, 89.335°E), covering an area of approximately 2,376 km² with a population of 2,909,624 [19]. The district receives an average annual rainfall of 1,872 mm, with temperatures ranging from 9.6°C to 36.8°C. The study was conducted in Santhia and Sujanagar upazilas, selected for their agricultural significance, across ten villages: Dhulauri, Gaurigram, Chitail, Khetupara, Sonatola, Satbaria, Harirampur, Bonkola, Char Tarapur, and Char Ramkantapur. Fig 1 presents the geographical location of the study area adapted from Banglapedia [18].

**Fig 1.**
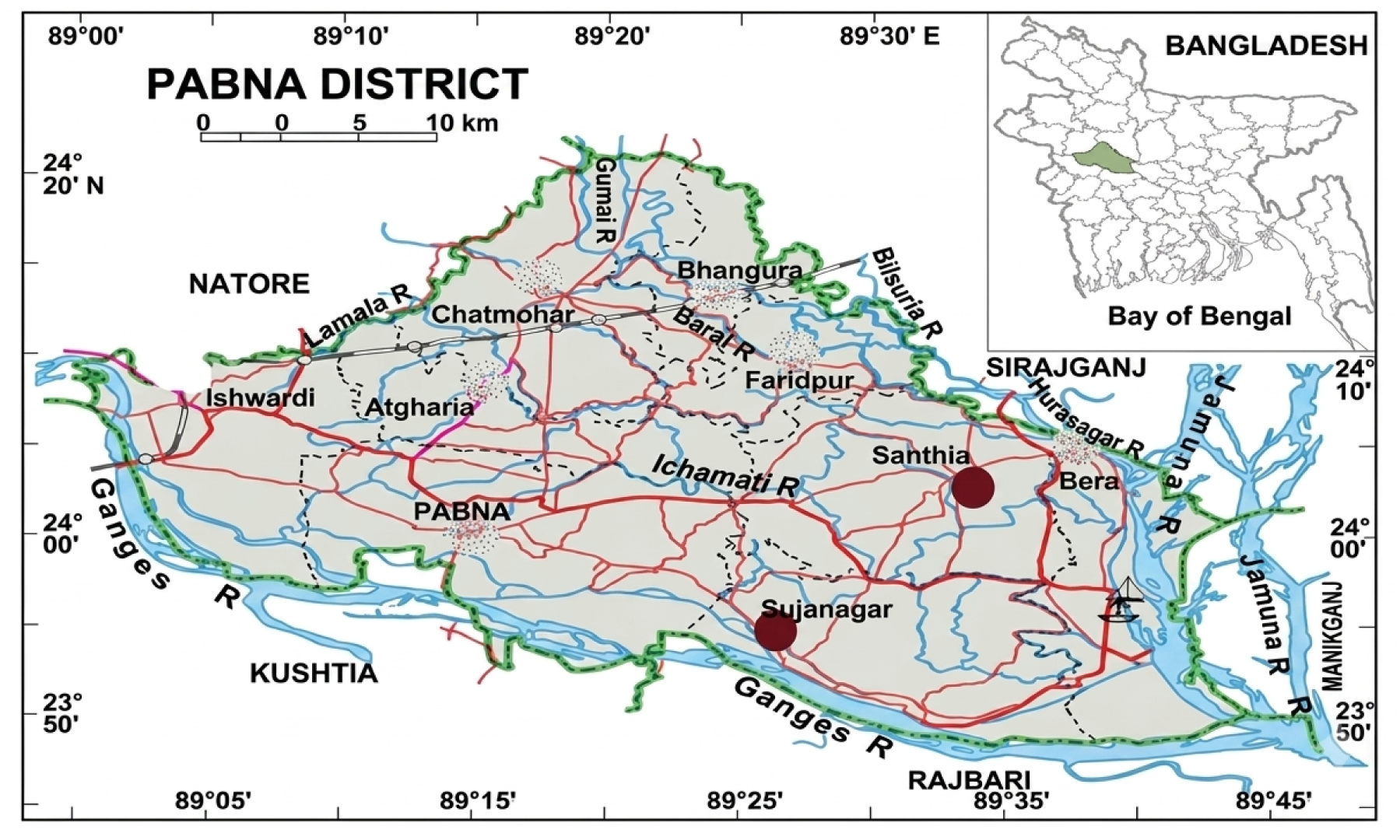
Map of Pabna District, Bangladesh. Red dots indicate Santhia and Sujanagar upazilas, where data collection was conducted. Adapted from Banglapedia [18].

### 2.2 Sampling procedure

The sampling frame was drawn from the farmer registry maintained by the local Department of Agricultural Extension (DAE) in the selected villages. Eligible farming households were selected using simple random (lottery) sampling, and one farmer per household was interviewed.

Inclusion criteria: respondents were smallholder farmers of any gender, actively engaged in crop production during the study period, able to understand Bangla, and willing to complete the survey.

Exclusion criteria: individuals not engaged in farm-level production (e.g., those involved solely in wholesale or retail trade), seasonal farmers, and those unable to complete the survey.

### 2.3 Sample size determination

The required sample size was calculated using the formula:

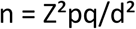

where Z = 1.96 (95% confidence level), p = 0.25 (estimated prevalence), q = 1−p, and d = 0.05 (margin of error), yielding a minimum required sample of 288 respondents. Accounting for an anticipated non-response rate, a final sample of 301 farmers was surveyed and included in the analysis.

### 2.4 Questionnaire development and data collection

A structured questionnaire comprising 40 closed and open-ended questions across seven sections was developed: (1) socio-demographic characteristics, (2) knowledge and adoption of environmentally sustainable farming practices, (3) soil testing and organic farming/manure use, (4) pesticide handling and management, (5) occupational health and safety practices, (6) post-harvest and market waste management, and (7) market sanitation and water facilities. The structure of the questionnaire is summarised in Fig 2. Each interview lasted approximately 40-45 minutes.

**Fig 2.**
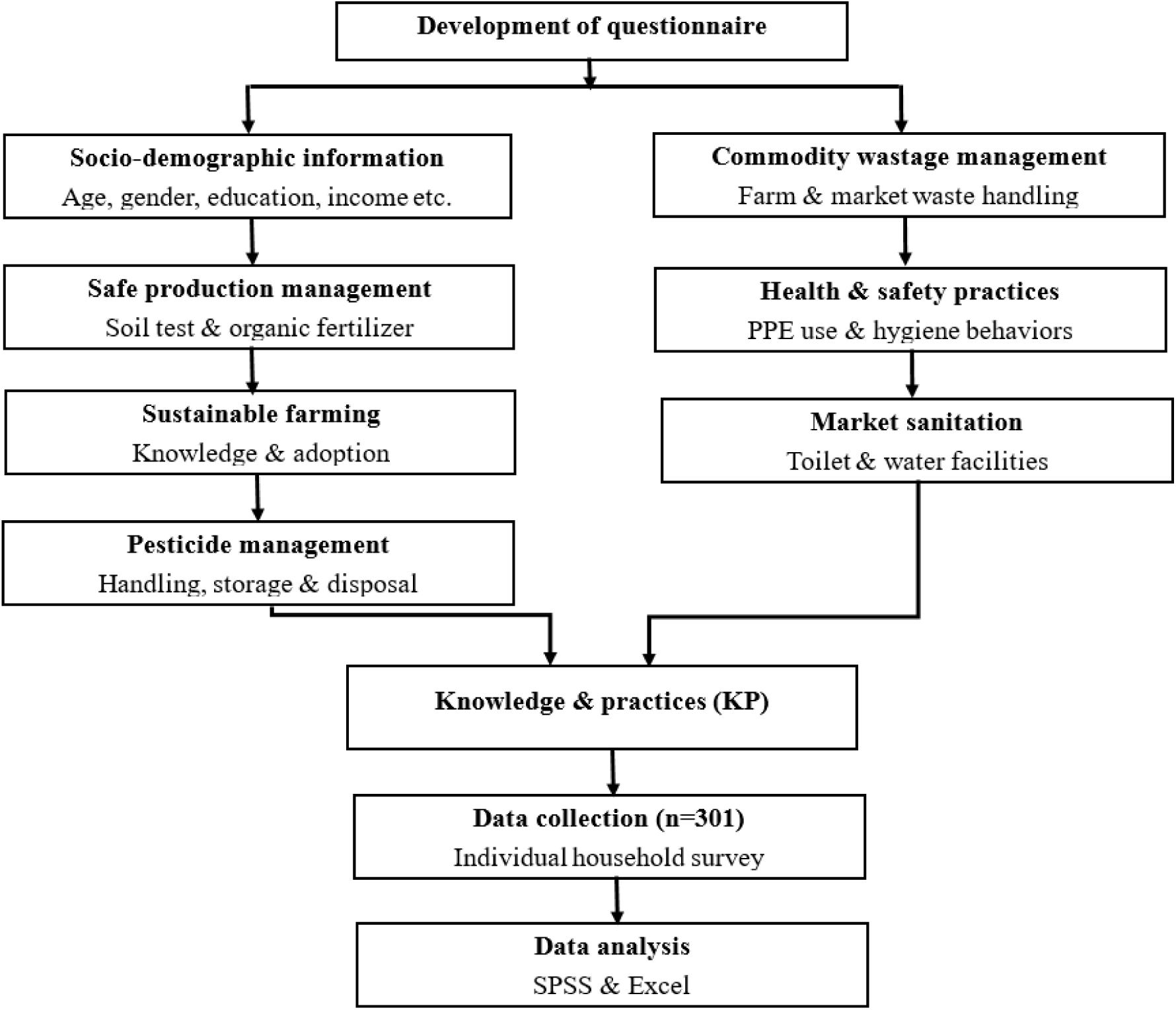
Structure of the study questionnaire, showing its seven content domains and the flow from questionnaire development to data analysis.

The questionnaire was originally developed in Bengali and translated into English. Prior to full data collection, it was pre-tested through key informant interviews and group discussions with 20 respondents, and revised accordingly. Ten trained enumerators, who completed a one-day training session covering item-by-item review and a field pilot test, conducted household interviews.

### 2.5 Data processing and statistical analysis

Completed questionnaires were reviewed on the day of collection, and any inconsistencies were resolved through follow-up phone contact with respondents. Data were coded and entered into International Business Machines (IBM) Statistical Package for the Social Sciences (SPSS) statistics version 20.0. Descriptive statistics (frequencies and percentages) were used to summarise socio-demographic characteristics and all knowledge- and practice-related variables. Summary outputs were subsequently organised in Microsoft Excel for the preparation of tables and figures presented in this manuscript.

### 2.6 Ethical considerations

Ethical approval to conduct this study was granted by the Mawlana Bhashani Science and Technology University’s Faculty of Life Science Ethical Review Committee in Tangail, Bangladesh. Before any data collection, researchers and enumerators obtained written consent from each participant, outlining the goals, risks, and benefits of participating in the study. For participants under the age of 18, written consent was additionally obtained from their guardian, alongside the participant’s own assent.

## 3. Results

### 3.1. Socio-demographic characteristics of respondents

Table 1 presents the demographic and socioeconomic characteristics of the surveyed farmers (N = 301). The majority of respondents were male (87.04%), while female respondents accounted for 12.96% of the sample. Regarding age distribution, most respondents (59.47%) were between 18 and 28 years old, followed by those aged 29-40 years (26.58%). Smaller proportions were over 40 years (9.97%) or under 18 years (3.99%). With respect to educational attainment, nearly two-thirds of respondents (63.46%) had no formal education. Among those with some education, 23.59% had completed primary level, 7.64% had graduated or held higher qualifications, and 5.32% had completed secondary or higher secondary education. Regarding monthly farm income, more than half of respondents (53.16%) earned less than 5,000 BDT per month, 37.87% earned between 5,000 and 10,000 BDT, and 8.97% earned more than 10,000 Bangladeshi Taka (BDT).

**Table 1:** Socioeconomic and demographic information of respondent households (N=301)

| Variable | n (%) |
| --- | --- |
| **Gender** |  |
| Male | 262(87.04) |
| Female | 39(12.96) |
| **Age (Years)** |  |
| <18 | 12(3.99) |
| 18-28 | 179(59.47) |
| 29-40 | 80(26.58) |
| >40 | 30(9.97) |
| **Level of Education** |  |
| Illiterate | 191(63.46) |
| Primary | 71(23.59) |
| Secondary/Higher secondary | 16(5.32) |
| **Monthly Income (BDT\*)** |  |
| <5000 | 160(53.16) |
| 5000-10000 | 114(37.87) |
| >10000 | 27(8.97) |

### 3.2. Knowledge, adoption, and perceived benefits of environmentally sustainable farming

Table 2 presents farmers’ knowledge and adoption of environmentally sustainable farming practices, along with the environmental and occupational health measures implemented on their farms. The largest proportion of respondents (39.87%) associated a healthy agricultural environment with soil, water, and organic matter, followed by land, crop, and market (33.55%); smaller proportions cited soil, fertiliser, and seed (16.94%), while 9.63% could not identify any components.

**Table 2.**
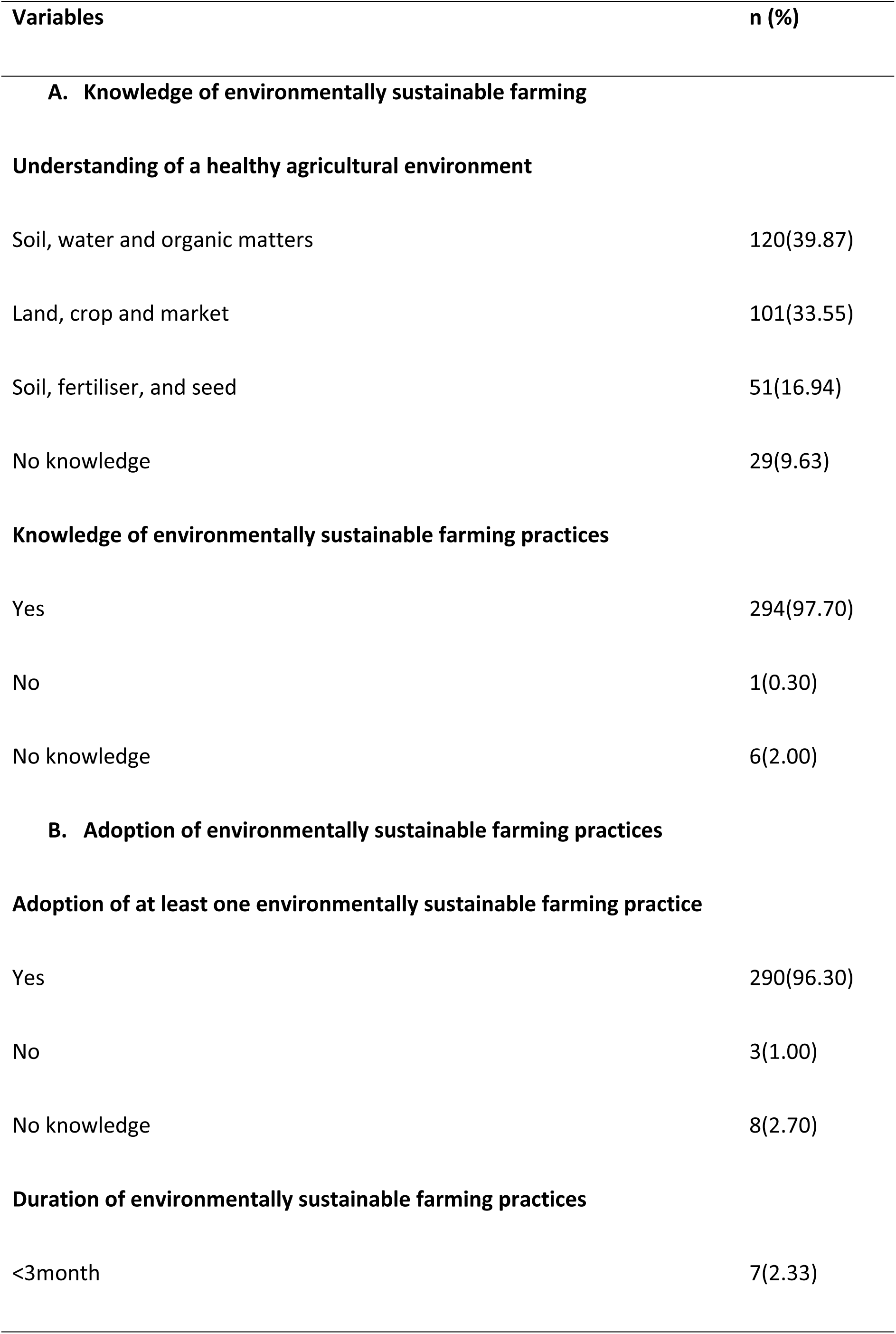
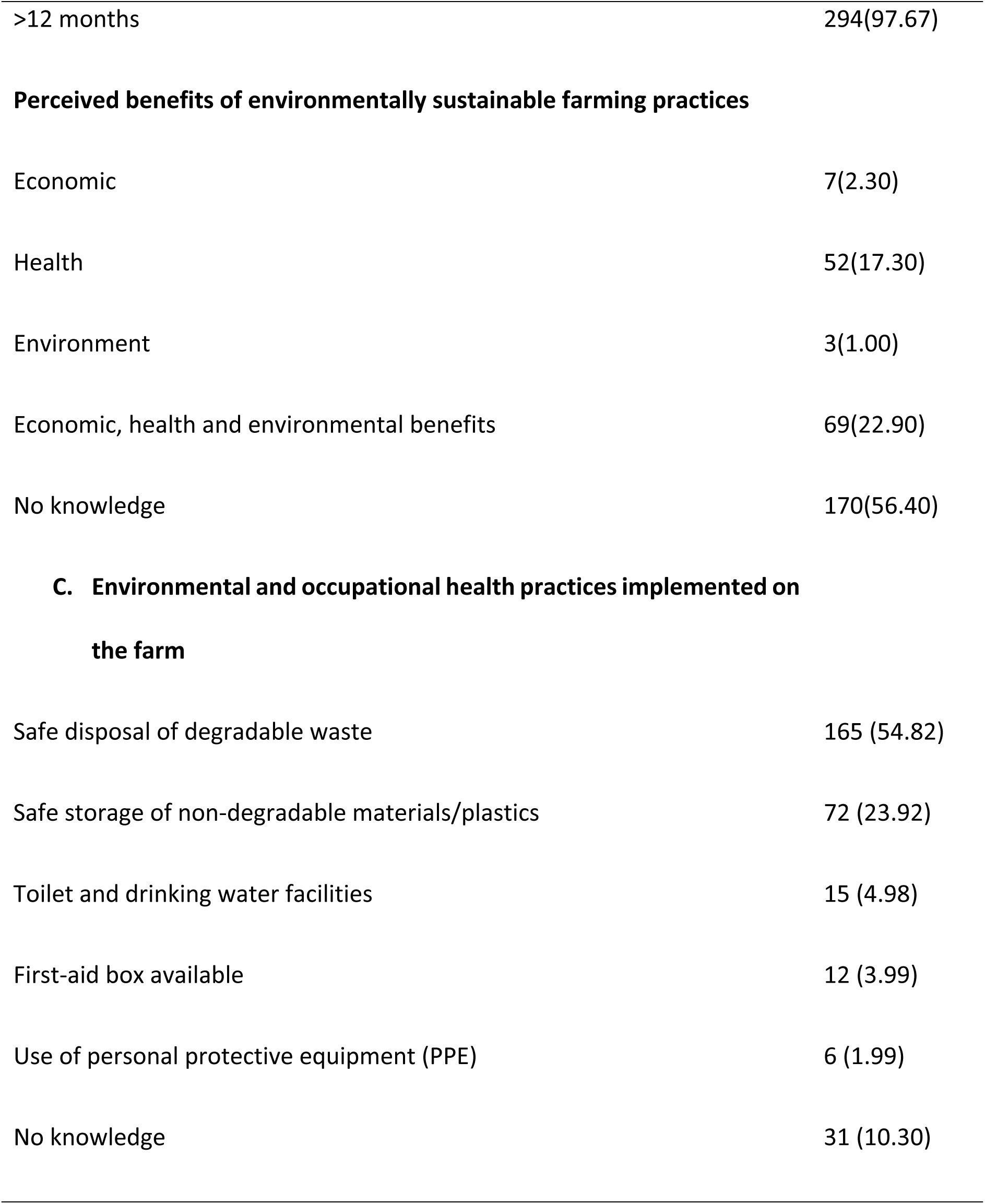
Farmers’ knowledge, adoption, and perceived benefits of environmentally sustainable farming practices (N=301)

| **Variables** | **n (%)** |
| --- | --- |
| **A. Knowledge of environmentally sustainable farming** |  |
| **Understanding of a healthy agricultural environment** |  |
| Soil, water and organic matters | 120(39.87) |
| Land, crop and market | 101(33.55) |
| Soil, fertiliser, and seed | 51(16.94) |
| No knowledge | 29(9.63) |
| **Knowledge of environmentally sustainable farming practices** |  |
| Yes | 294(97.70) |
| No | 1(0.30) |
| No knowledge | 6(2.00) |
| **B. Adoption of environmentally sustainable farming practices** |  |
| **Adoption of at least one environmentally sustainable farming practice** |  |
| Yes | 290(96.30) |
| No | 3(1.00) |
| No knowledge | 8(2.70) |
| **Duration of environmentally sustainable farming practices** |  |
| <3month | 7(2.33) |
| >12 months | 294(97.67) |
| **Perceived benefits of environmentally sustainable farming practices** |  |
| Economic | 7(2.30) |
| Health | 52(17.30) |
| Environment | 3(1.00) |
| Economic, health and environmental benefits | 69(22.90) |
| No knowledge | 170(56.40) |
| **C. Environmental and occupational health practices implemented on the farm** |  |
| Safe disposal of degradable waste | 165 (54.82) |
| Safe storage of non-degradable materials/plastics | 72 (23.92) |
| Toilet and drinking water facilities | 15 (4.98) |
| First-aid box available | 12 (3.99) |
| Use of personal protective equipment (PPE) | 6 (1.99) |
| No knowledge | 31 (10.30) |

Almost all respondents (97.70%) reported knowledge of environmentally sustainable farming practices, and a similarly high proportion (96.30%) reported adopting at least one such practice, with the vast majority of adopters (97.67%) having done so for more than 12 months. Despite this high reported awareness and adoption, more than half of respondents (56.40%) could not identify any specific benefit of sustainable practices; among those who could, 22.90% identified combined economic, health, and environmental benefits.

With respect to on-farm practices, safe disposal of degradable waste was the most commonly reported measure (54.82%), followed by safe storage of non-degradable materials and plastics (23.92%). Practices more directly related to occupational health and safety were far less common, toilet and drinking water facilities (4.98%), first-aid availability (3.99%), and PPE use (1.99%), the last being the lowest of all reported practices.

### 3.3. Soil testing, organic farming knowledge, and manure use

Table 3 presents farmers’ knowledge and practices related to soil testing, organic farming, and manure use. Knowledge of safe or certified commodity production standards was reported by very few respondents (2.30%). Just over a quarter (27.57%) reported conducting soil tests, and among these, 28.92% reported benefiting from soil testing before planting.

**Table 3:**
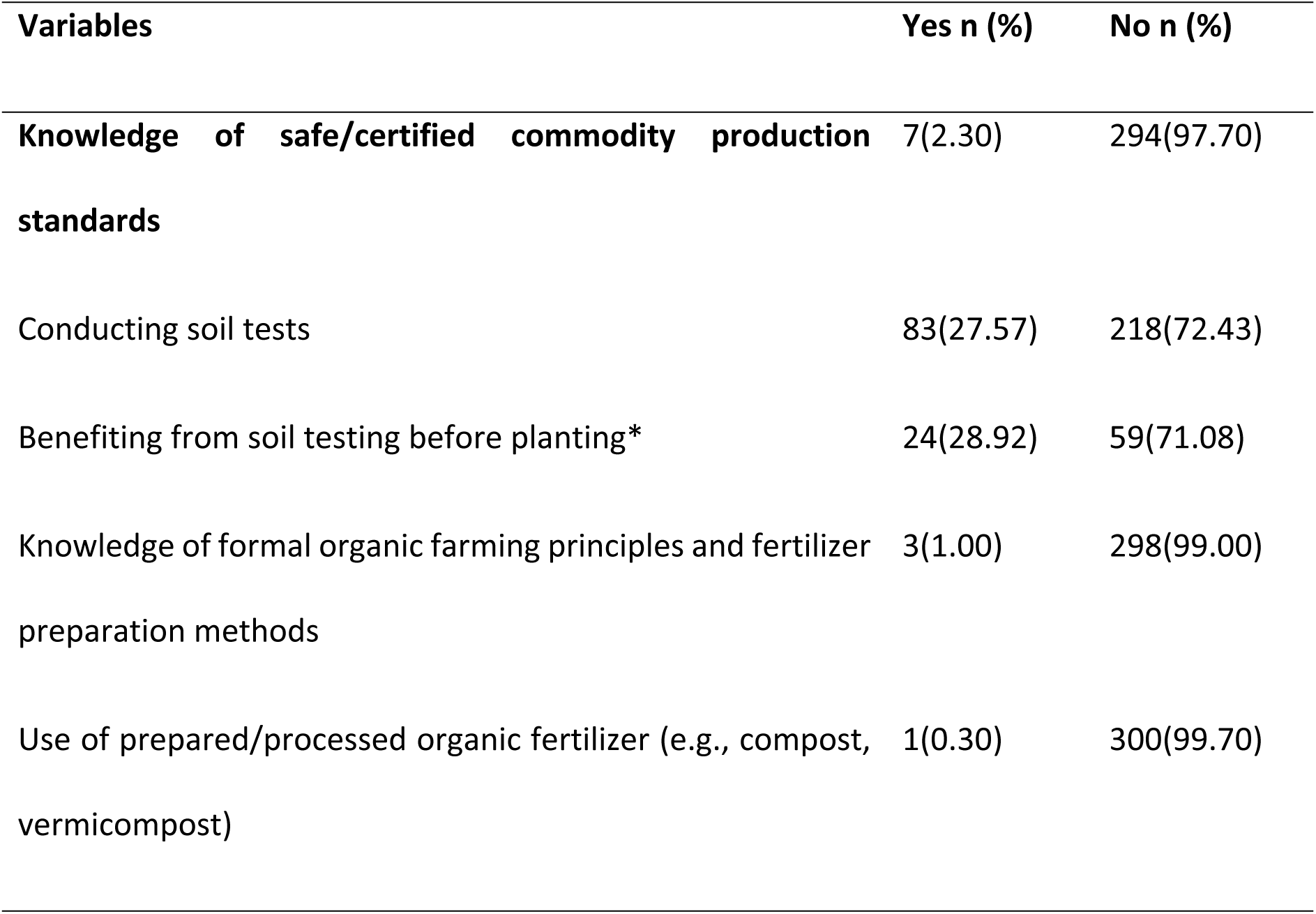
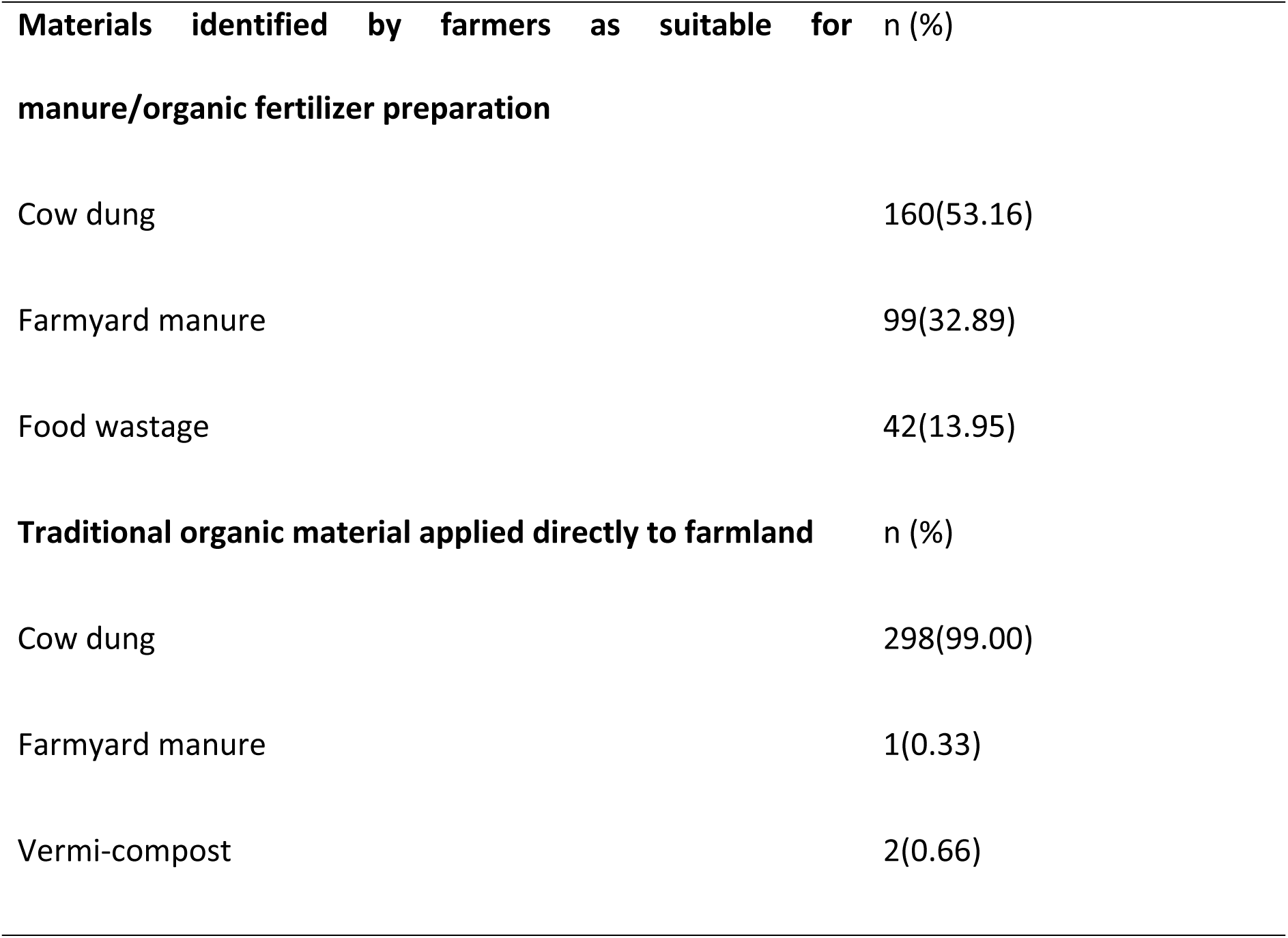
Farmers’ knowledge and practices related to soil testing, organic farming, and manure use (N = 301)

| Variables | Yes n (%) | No n (%) |
| --- | --- | --- |
| **Knowledge of safe/certified commodity production standards** | 7(2.30) | 294(97.70) |
| Conducting soil tests | 83(27.57) | 218(72.43) |
| Benefiting from soil testing before planting* | 24(28.92) | 59(71.08) |
| Knowledge of formal organic farming principles and fertilizer preparation methods | 3(1.00) | 298(99.00) |
| Use of prepared/processed organic fertilizer (e.g., compost, vermicompost) | 1(0.30) | 300(99.70) |

| **Materials identified by farmers as suitable for** |  | **n (%)** |
| --- | --- | --- |
| **manure/organic fertilizer preparation** |  |  |
| Cow dung |  | 160(53.16) |
| Farmyard manure |  | 99(32.89) |
| Food wastage |  | 42(13.95) |
| **Traditional organic material applied directly to farmland** |  | **n (%)** |
| Cow dung |  | 298(99.00) |
| Farmyard manure |  | 1(0.33) |
| Vermi-compost |  | 2(0.66) |

Formal knowledge of organic farming principles was reported by only 1.00% of respondents, and use of prepared or processed organic fertilizer (e.g., compost, vermicompost) was reported by a single farmer (0.30%). When asked which materials they considered suitable for manure preparation, cow dung was most commonly identified (53.16%), followed by farmyard manure (32.89%) and food waste (13.95%). In terms of material actually applied directly to farmland, cow dung was reported by the vast majority (99.00%), while farmyard manure (0.33%) and vermicompost (0.66%) were rarely used.

### 3.4. Pesticide knowledge, handling, and occupational health practices

Table 4 presents farmers’ knowledge and safety practices related to pesticide handling and application. Almost all respondents reported understanding pesticide application according to label instructions (98.00%) and colour coding (99.34%). Nearly three-quarters (73.09%) were aware that pesticides can harm beneficial organisms, yet a similar proportion (73.75%) reported using a mixture of multiple pesticides during application.

**Table 4:** Farmers’ knowledge and safety practices related to pesticide handling and application (N=301)

| Variables | Yes n (%) | No n (%) |
| --- | --- | --- |
| A. Knowledge of pesticide use |  |  |
| Knowledge of pesticide application according to label instructions | 298(98.00) | 6(2.00) |
| Knowledge of pesticide colour coding | 299(99.34) | 3(0.66) |
| Use multiple pesticides mixture for application | 222(73.75) | 79(26.25) |
| Knowledge that pesticides destroy beneficial organisms | 220(73.09) | 81(26.91) |
| B. Pesticide handling and management practices | n (%) |  |
| Source of pesticide information |  |  |
| Pesticide dealer | 160(53.16) |  |
| Pesticide company | 4(1.33) |  |
| Neighborhood farmers | 106(35.22) |  |
| Agriculture extension office | 15(4.98) |  |
| Others* | 16(5.32) |  |
| **Timing of pesticide application** |  |  |
| Morning | 7(2.33) |  |
| Midday | 2(0.66) |  |
| Evening | 292(97.01) |  |
| **Disposal/storage of empty pesticide containers** |  |  |
| Land's side | 3(1.00) |  |
| Road | 3(1.00) |  |
| Land's hole | 258(85.71) |  |
| Other places + | 37(12.29) |  |
| **C. Occupational health and safety practices** | **Yes n (%)** | **No n (%)** |
| Smoking during the spraying of Pesticide | 221(73.42) | 80(26.58) |
| Cleaning spray equipment after use | 299(99.30) | 2(00.70) |
| Knowledge of PPE use | 5(1.66) | 296(98.34) |
| Wearing at least one PPE | 32(10.63) | 269(89.37) |
| Wearing a mask | 4(1.33) | 297(98.67) |
| Washing with soap after spraying | 67(22.26) | 234(77.74) |

Pesticide dealers were the primary source of information (53.16%), followed by neighbouring farmers (35.22%); few relied on extension officers (4.98%) or pesticide companies (1.33%). Most respondents applied pesticides in the evening (97.01%), and the majority (85.71%) disposed of empty containers by burying them in a hole near their land.

Occupational health and safety practices were generally poor. While nearly all respondents reported cleaning spray equipment after use (99.30%), very few reported knowledge of PPE (1.66%) or PPE use (10.63%), and mask use was particularly low (1.33%). Only 22.26% reported washing with soap after spraying, and nearly three-quarters (73.42%) reported smoking during application.

### 3.5. Post-harvest waste management and market sanitation

Table 5 presents farmers’ post-harvest waste management practices and market sanitation conditions. Most respondents (88.70%) reported managing commodity waste after harvesting; among these, piling waste at a single farm site was most common (74.75%), followed by burning and mixing residues into soil (23.92%), while composting was rare (1.33%).

**Table 5.**
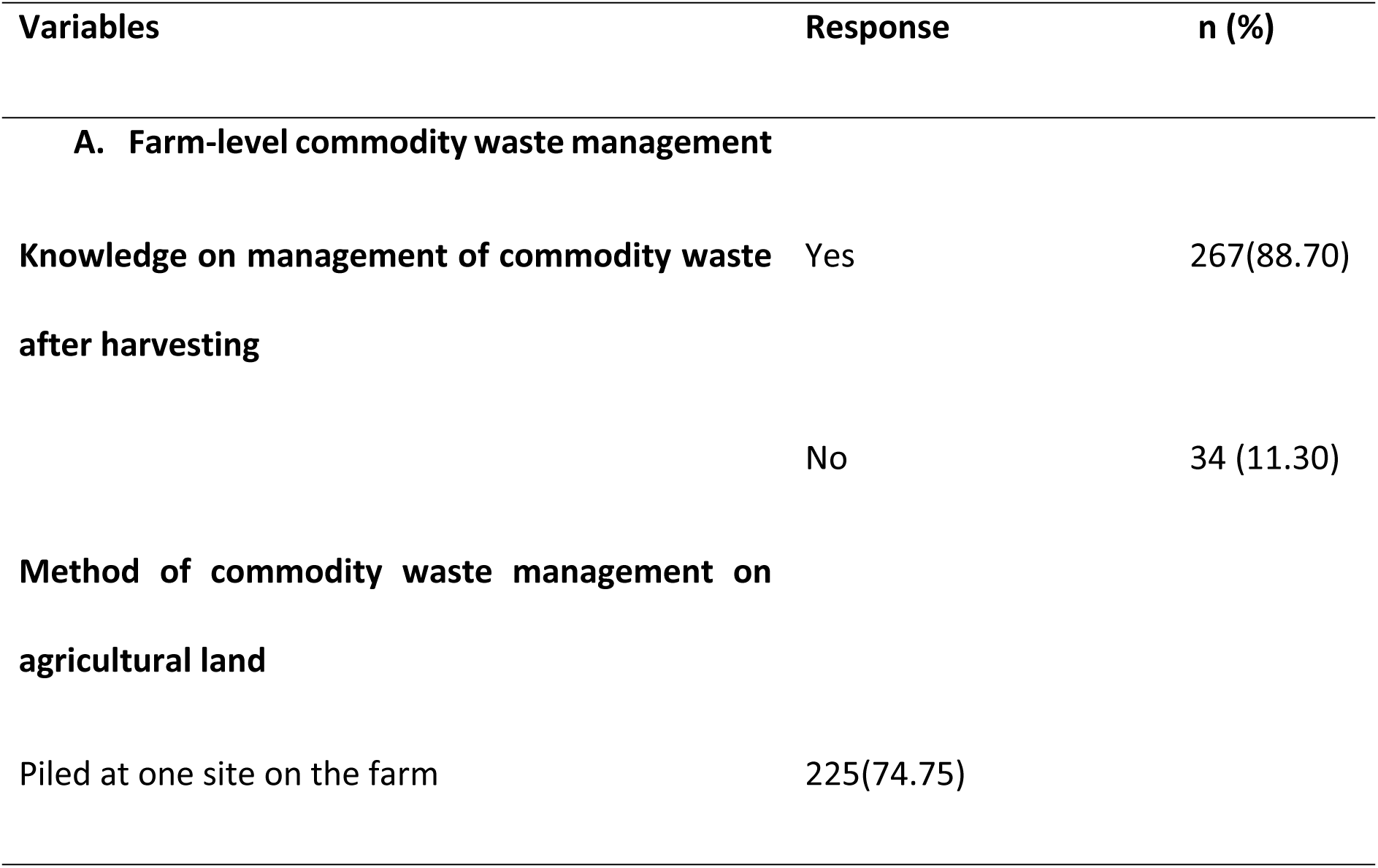
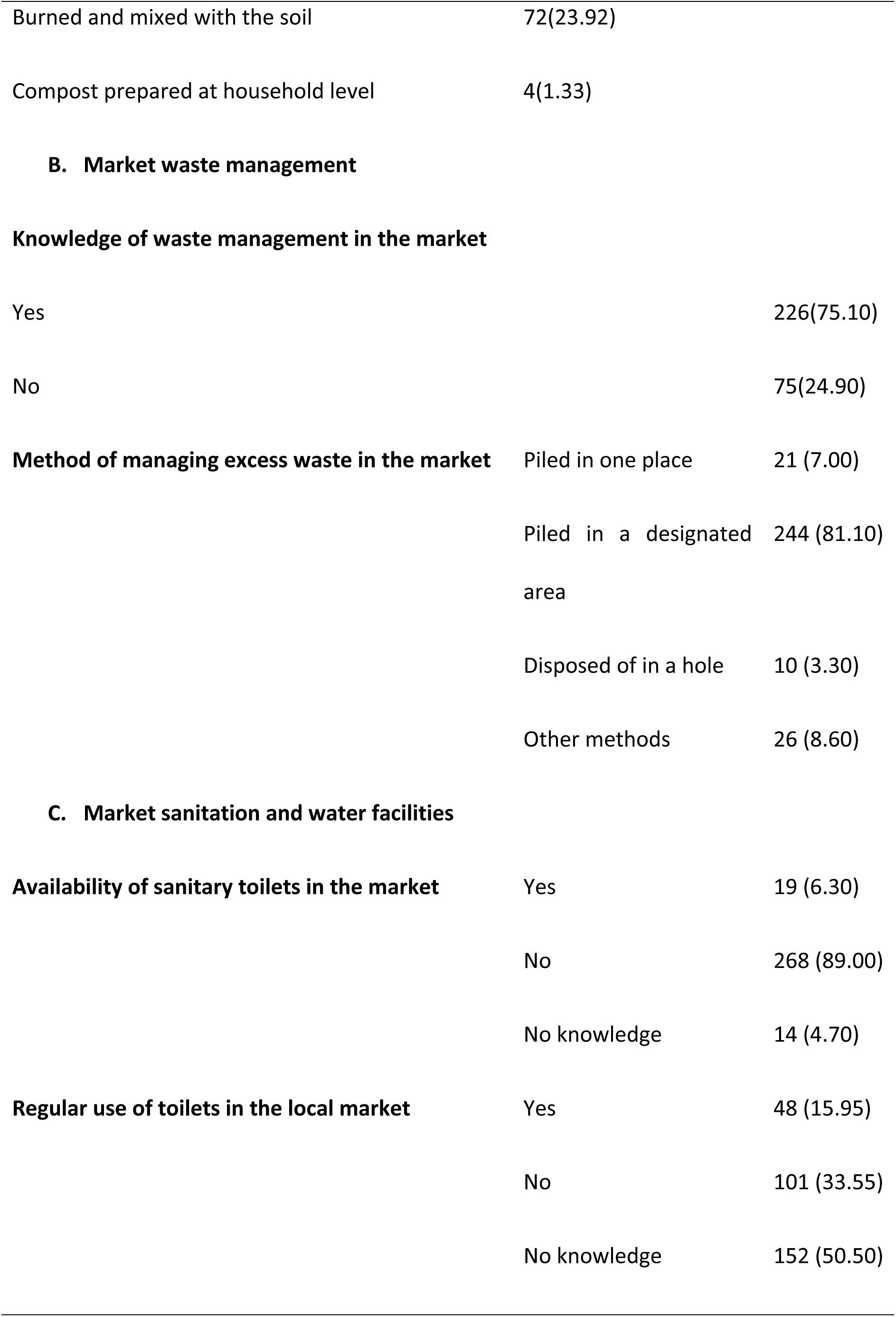
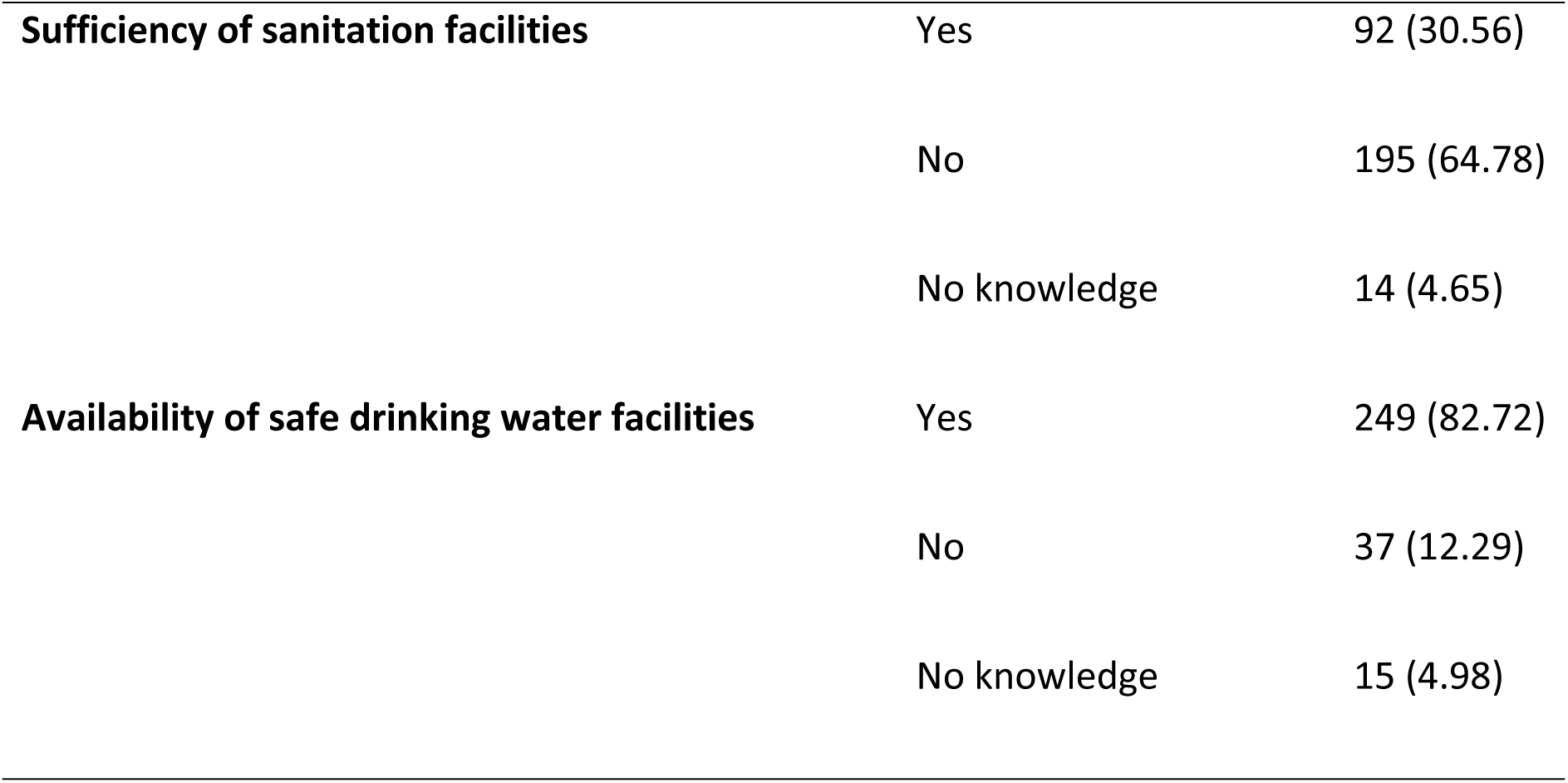
Post-harvest waste management and market sanitation practices among farmers (N=301)

At the market level, most respondents (75.10%) reported awareness of waste management practices, and most of those managing waste used designated disposal areas (81.10%). Sanitation infrastructure, however, was inadequate: sanitary toilets were unavailable according to 89.00% of respondents, only 15.95% reported regular toilet use, and 64.78% considered facilities insufficient. In contrast, safe drinking water was reported as available by most respondents (82.72%).

### 3.6. Self-reported health symptoms following pesticide application

Fig 3 presents farmers’ self-reported acute health symptoms following pesticide application. Eye irritation or burning was by far the most common symptom, affecting 92.69% of respondents, followed by physical weakness (4.32%), headache (1.66%), and vomiting (1.00%).

**Fig 3.**
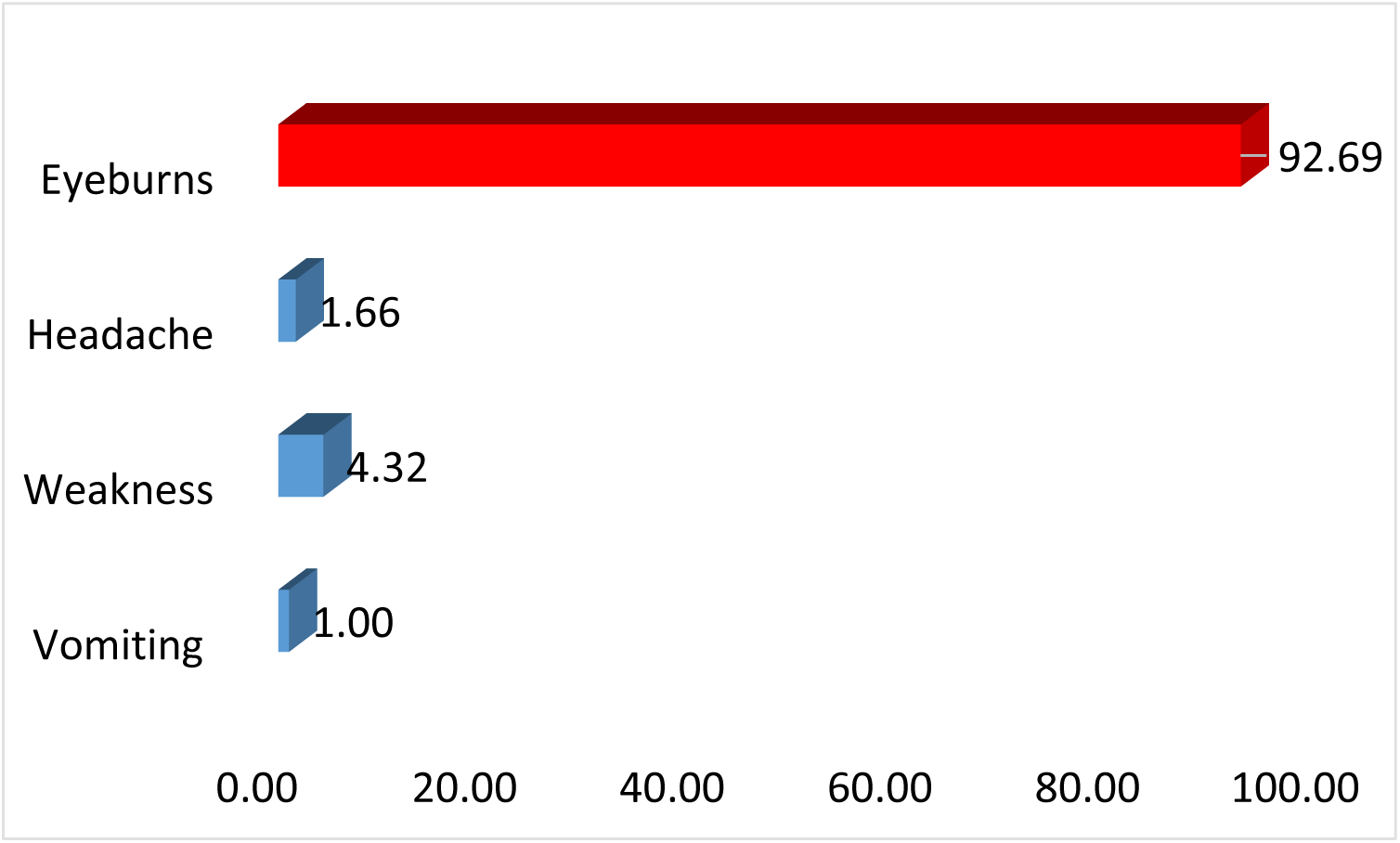
Self-reported health problems associated with pesticide exposure among surveyed farmers.

## 4. Discussion

### 4.1. Socio-demographic characteristics

The socioeconomic profile of the surveyed farmers provides important context for understanding occupational health and safety (OHS) practices in Bangladesh’s agri-food system. The predominance of male respondents reflects the traditional division of labour in Bangladeshi agriculture, where field operations and pesticide application are mainly carried out by men, though the presence of female respondents highlights the need to include women in future OHS training.

The concentration of respondents in the 18-28-year age group indicates a predominantly young workforce. Farmers’ age has been linked to protective behaviour and safe pesticide handling in several studies [20, 21, 22], although the relationship between age and PPE use is inconsistent across the literature; some report a positive association [23], while others report older farmers relying on traditional methods and using PPE less [24].

Educational attainment was low overall, with nearly two-thirds of respondents having no formal education. This is consistent with previous studies linking education to awareness and use of protective measures [25, 26, 27, 28, 29, 30], and with evidence identifying education as the strongest predictor of PPE use among farmers, noting that poorly educated farmers face greater exposure risk due to difficulty understanding usage instructions [31, 24].

Economic constraints further compound these risks: more than half of respondents earned less than 5,000 BDT monthly, which may limit their capacity to purchase PPE or adopt safer practices, consistent with previous studies [32, 33]. Overall, age, education, and income emerge as key factors shaping OHS practices among farmers.

### 4.2. Sustainable farming awareness versus practice

Although almost all respondents reported knowledge of sustainable farming practices and adoption of at least one such practice, more than half were unable to identify any specific benefit. This pattern is consistent with self-reported awareness measures more broadly, which may capture general familiarity rather than in-depth technical understanding. The relatively low educational attainment of the respondents should also be considered when interpreting these findings, and the reported levels of awareness and adoption should therefore be regarded as indicative rather than comprehensive.

A further concern is the disparity between environmental and occupational health practices. While waste-related measures were relatively common, practices directly protecting worker health were reported by fewer than 5% of respondents, suggesting environmental sustainability and occupational safety are not yet equally prioritised. This study assessed waste disposal in broad terms, whereas existing literature often focuses specifically on pesticide storage; for example, most farmers in one study stored pesticides in designated chemical stores, while others used open sheds or fields [11], a more specific framing that may capture risk behaviours a general ‘waste disposal’ measure does not.

The low adoption of occupational health measures aligns with evidence linking safety behaviour to awareness and training, while favourable attitudes toward ecological agriculture have been associated with older age, higher education, and greater extension contact [34].

As this study’s respondents were predominantly young and had low educational attainment, these findings may partly explain the observed gap between general awareness and limited adoption of occupational safety measures.

### 4.3. Soil testing and organic farming knowledge

Overall knowledge of safe commodity production standards and formal organic farming principles was very low, despite near-universal application of cow dung as a soil input. This suggests that farmers use cow dung mainly as a traditional farming practice rather than as part of an organic farming approach. Similar findings have been reported in previous studies, where most farmers ranked cow dung and poultry manure among the most important practices for maintaining soil health despite having limited knowledge of organic farming principles [15].

The low uptake of soil testing is consistent with wider evidence of low adoption of soil testing and fertilizer recommendation services in Bangladesh, where adoption has been found to be strongly shaped by farmers’ age, education, and income [13], and where only moderate adoption of recommended soil fertility practices has been observed even where awareness exists [14]. The limited shift from raw manure application toward structured organic fertilizer preparation mirrors barriers identified by Hasan et al. (2025), who reported that limited access to information, inadequate training, and low awareness of proper preparation methods similarly constrained biofertilizer adoption among farmers in northern Bangladesh [16].

### 4.4. Pesticide knowledge, handling practices, and occupational safety

Findings indicate that farmers’ knowledge of pesticide labels and hazard information has not translated into safer practices. Despite widespread awareness that pesticides can harm beneficial organisms, most respondents still reported mixing multiple pesticides to boost yield. This finding is consistent with evidence that farmers may adopt risky practices despite understanding the hazards, often due to limited practical training or greater concern for economic returns [35, 36].

The heavy reliance on pesticide dealers as the primary information source is a further concern, as retailers may lack technical training or have financial incentives to downplay health risks [37]. This finding is in line with Sankoh et al. (2016), who similarly reported that over half of surveyed rice farmers in Sierra Leone depended on vendors for pesticide information [26]. The widespread practice of burying empty containers near farmland raises concern about soil and water contamination, consistent with risks documented by Atreya et al. (2012) among vegetable farmers in Nepal [38].

OHS practices were markedly poor. Almost no respondents reported knowledge or use of PPE, aligning with evidence that the majority of farmers elsewhere similarly did not use PPE when handling pesticides [11]. Combined with low rates of post-application hand washing and high rates of smoking during application, these findings point to substantial gaps in protective behaviour. PPE use has been associated with a 44-80% reduction in poisoning [39], while its absence increases dermal and respiratory exposure [40]. These results highlight an urgent need for practical, hands-on training in protective practices, rather than general awareness-building alone.

### 4.5. Post-harvest waste management and market sanitation

Although most farmers reported managing commodity waste after harvest, the methods used were basic. Piling waste at a single farm site was most common, while burning and mixing residues into soil was reported by nearly a quarter of respondents. This practice may contribute to air pollution and reduced soil quality over time. Composting was rarely used, suggesting limited awareness of or access to composting methods. Extension services could help promote low-cost composting techniques, consistent with evidence that extension support plays an important role in helping smallholder farmers adopt better post-harvest management practices [41].

At the market level, awareness of waste management was not matched by adequate sanitation infrastructure, suggesting the problem is structural rather than knowledge-based. This is consistent with evidence that poor market infrastructure and sanitation are common barriers to food safety in informal markets in low and middle-income countries [17]. The relatively good availability of safe drinking water is a positive finding, though alone insufficient to ensure hygienic market conditions.

### 4.6. Health symptoms following pesticide exposure

The overwhelming predominance of eye irritation in this study contrasts with more evenly distributed symptom profiles reported elsewhere, where headache, itchy eyes, skin irritation, and fatigue were all reported at comparably high rates [11]. This difference may reflect the specific exposure routes and protective gaps identified in this study. Mask and PPE use overall were minimal, leaving the eyes among the least protected body parts during spraying.

Beyond acute symptoms, pesticide exposure has been linked in the broader literature to respiratory, dermatological, gastrointestinal, and reproductive disorders [6, 26], as well as long-term neurological and carcinogenic risks under chronic exposure [5]. Agricultural workers more broadly have also been reported to experience skin and eye complications linked to pesticide handling, particularly where hot and humid conditions discourage consistent use of protective equipment [42]. These risks should be understood within the broader context of occupational health among smallholder farmers, who are commonly exposed to multiple concurrent stressors that collectively affect physical health, productivity, and quality of life [43]. Although farmers in this study generally recognised the productivity benefits of pesticide use, this awareness has not translated into consistent adoption of safer handling practices.

### 4.7. Overall implications

These findings indicate that farmers in this study possess reasonably good factual knowledge of pesticide labelling, sustainable farming concepts, and waste management, but this knowledge has not consistently translated into protective behaviour, particularly regarding PPE use, safe pesticide disposal, and market sanitation. This pattern is consistent with findings from Indonesia, where farmers similarly demonstrated good knowledge of good agricultural practices (GAP) for safe pesticide usage, yet showed significantly lower levels of actual application, especially regarding the use of gloves and masks and the proper disposal of pesticide containers [12]. Age, education, and income emerge as recurring structural factors underlying this knowledge-practice gap in the present study.

Strengthening extension services with a specific focus on practical, hands-on occupational safety training, rather than general awareness-building alone, is recommended. Targeted infrastructure investment in market sanitation may further help close this gap and support both farmer wellbeing and the sustainable transformation of Bangladesh’s agri-food system.

## 5. Conclusion and recommendations

This study set out to assess farmers’ knowledge and practices related to occupational health, pesticide handling, environmentally sustainable farming, and market sanitation in Pabna District, Bangladesh. It can be concluded from the results of this study that farmers generally possess reasonable factual knowledge across these domains, most were aware of pesticide label instructions, understood the concept of a healthy agricultural environment, and recognised the general risks associated with pesticide exposure. However, this knowledge did not consistently translate into safe or sustainable practice. This gap was most pronounced in occupational health and safety behaviours. Despite near-universal awareness of pesticide hazards, fewer than 11% of farmers reported using any personal protective equipment, and mask use was almost entirely absent. Correspondingly, eye irritation was reported by the vast majority of farmers following pesticide application, underscoring the practical health consequences of this knowledge-practice gap.

Similar patterns were observed beyond pesticide handling. Farmers frequently relied on cow dung as a soil input without a structured understanding of organic farming principles, and the uptake of soil testing services remained low despite their availability. At the market level, awareness of waste management practices was not matched by adequate sanitation infrastructure, with the large majority of markets lacking sanitary toilets.

Therefore, it is recommended that farmer training in Pabna District should move beyond general awareness-building toward practical, hands-on instruction, particularly in the use of personal protective equipment, safe pesticide disposal, and structured organic fertilizer preparation. As pesticide dealers were the primary source of information for farmers, training programmes should be designed not only for farmers but also for pesticide retailers, in order to promote accurate guidance on the safe use of pesticides. At the market level, addressing the sanitation gap will likely require direct infrastructure investment such as the construction and maintenance of sanitary toilets rather than farmer-education efforts alone. Delivering this training through participatory approaches, such as farmer field schools and demonstration plots, may improve the likelihood that farmers translate existing knowledge into safer, more sustainable practice.

## 6. Strengths and limitations of the study

The main strength of this study is its integrated assessment of farmers’ knowledge and practices across multiple stages of the agricultural value chain, from pre-harvest soil and pesticide management through to post-harvest waste handling and market sanitation within a single farming population in Pabna District. This breadth distinguishes it from much of the existing literature, which typically examines a single domain, such as pesticide safety alone, in isolation.

However, several limitations should be acknowledged. First, the sample was drawn from farmers in two upazilas of Pabna District, and findings may not be generalisable to farmers in other regions of Bangladesh with different cropping patterns or socio-economic conditions. Second, all practice-related data were based on farmers’ self-reports rather than direct observation, which may be subject to recall bias or social desirability bias, particularly for behaviours farmers know are considered unsafe. Third, the cross-sectional design captures a single point in time and cannot establish causal relationships between socio-demographic characteristics and reported practices. Finally, this study relied on descriptive statistics, future research could apply inferential statistical methods to formally test associations between farmers’ characteristics and their knowledge or practice scores.

## Declarations

### Author contribution statement

Shahnaj Parvin: Conceptualization, Data Curation, Methodology, Formal analysis, Visualization, Validation, Supervision, Project administration, Writing - original draft preparation, Writing - review & editing.

Md. Obaidur Rashid: Conceptualization, Investigation, Methodology, Formal analysis, Validation, Data curation, Supervision, Software, Writing - original draft, Writing - review & editing.

Md. Maksudur Rahman: Investigation, Supervision, Validation, Visualization, Writing - review & editing.

Abdullah Al Zaber: Formal analysis, Resources, Investigation, Visualization, Writing - review & editing, Validation.

Md. Bakhtiar Hossain: Investigation, Data curation, Resources, Validation, Writing - review & editing.

Rokeya Begum: Conceptualization, Methodology, Formal Analysis, Supervision, Validation, Resources, Writing - review & editing.

Md. Azizul Haque: Conceptualization, Methodology, Supervision, Validation, Resources, Writing - review & editing.

All authors have read and approved the final version of the manuscript and agree to be accountable for all aspects of the work, ensuring that questions related to the accuracy or integrity of any part of the work are appropriately investigated and resolved.

### Funding statement

No grants, financial support, or other funding were received from any public, private, commercial, social, governmental, or non-profit funding agency for the conduct of this study.

### Data availability statement

Data will be made available on request.

### Declaration of interests statement

The authors declare no conflict of interest

## Acknowledgements

The authors sincerely thank all the farmers of Pabna District who generously participated in this study and contributed their time and insights. We are especially grateful to the students of Mawlana Bhashani Science and Technology University (MBSTU) who supported the data collection process in the field, particularly Ayesha Siddika, Research Assistant, for her valuable support during data collection and field visits.

We are also grateful to Rakibul Hasan for supervisory guidance and constructive feedback throughout the study, and to Robiul Islam for statistical advice and assistance with data analysis.

We further extend our thanks to the District Agriculture Office, the Food Safety Office, and the Civil Surgeon’s Office of Pabna for their valuable technical guidance and cooperation throughout the study. We also acknowledge the faculty members and staff of the Faculty of Life Science, and the Department of Food Technology and Nutritional Science, Mawlana Bhashani Science and Technology University, for their institutional support, guidance, and assistance in refining the English language of this manuscript.

## Notes

### Competing Interest Statement

The authors have declared no competing interest.

### Author Declarations

The study was reviewed and approved by the Department of Food Technology and Nutritional Science Ethics Committee, Mawlanan Bhashani Science and Technology University, Bangladesh. Approval No. FN/FW/Internal/03/2024 Date: 03 March 2024.

